# Interconnected Challenges in Dementia Caregiving: A Co-occurrence Network Analysis of Burden, Unmet Needs, and System Failures Among Caregivers

**DOI:** 10.64898/2026.08.12.26360253

**Authors:** Yeon Mi Hwang, Tushar Mungle, Audrey Angel Kwan, Malvika Pillai, Michelle Sahai, Madelena Ng, Rebecca Handler, Tina Hernandez-Boussard

**Author notes:** Correspondence to: Dr. Yeon Mi Hwang, PhD, Department of Medicine, Stanford University, 3180 Porter Drive, Palo Alto, CA, 94304, Dr. Tushar Mungle, PhD, Department of Medicine, Stanford University, 3180 Porter Drive, Palo Alto, CA, 94304. Equal contribution (Yeon Mi Hwang, Tushar Mungle, Audrey Angel Kwan).

## Abstract

**Background:** Alzheimer’s Disease and Related Dementias (ADRD) is a growing global public health challenge, and caregivers experience high rates of burden, unmet needs, and system failures. These challenges vary by caregiver role and relationship to the care recipient, reflecting the heterogeneous nature of caregiving. Yet prior work has largely studied burden, unmet needs, and system failures as separate domains rather than examining how they co-occur within individual caregivers.

**Methods:** We applied an LLM-based classification framework (Claude 3.5 Sonnet) to 7,198 posts from three ALZConnected caregiver forums (general, spouse/partner, and adult child caregivers), coding each post for burden, unmet needs, and system failures across 9, 12, and 10 categories respectively. We compared expression rates by caregiver role (primary vs. secondary) and relationship to the care recipient (spousal vs. child) and used post-level co-occurrence networks to map how categories cluster within and across domains.

**Results:** Burden was expressed in 89.0% of posts and unmet needs in 93.3%, while system failures appeared in 34.8%. Primary caregivers reported burden more often than secondary caregivers (91.6% vs. 84.7%), while secondary caregivers reported more unmet needs (94.6% vs. 92.5%) and more system failures (37.2% vs. 33.4%). Child caregivers reported higher rates than spousal caregivers across all three domains. Co-occurrence networks showed dense within-domain clustering (density 0.61-0.65) and 84 significant cross-domain connections, with the strongest links between behavioral/safety burden and safety-management needs (21.7% of posts) and between emotional burden and emotional-support needs (20.9%).

**Conclusion:** Burden, unmet needs, and system failures are not independent problems but form interconnected challenge ecosystems that vary by caregiver role and relationship. This suggests caregiver support should be designed around these connected patterns rather than treated as separate, single-domain interventions.

## 1. Introduction

Alzheimer’s disease and related dementias (ADRD) are among the most pressing public health challenges facing aging societies worldwide. An estimated 57 million people are currently affected by ADRD and this number is projected to increase to 153 million by 2050^1^. ADRD places increasing demands not only on individuals living with the disease but on the families and health systems that support them. In the United States (US), more than 11 million family members and other unpaid caregivers provided an estimated 18.4 billion hours of care, valued at $346.6 billion to people with ADRD in 2023^2^. When formal medical and long-term care costs are included, the total economic burden of ADRD reaches $781 billion annually in the US, with Medicare and Medicaid covering 20% of that cost.^3^ Although these programs partially offset formal care expenses, caregivers still bear substantial financial, time, and well-being challenges. They experience higher rates of depression, anxiety, social isolation, and physical health decline than both non-caregivers and caregivers of persons with other conditions.^4,5^ As dementia prevalence continues to rise, understanding and addressing caregiver challenges has become an increasingly urgent public health priority.

Prior research has identified several dimensions of caregiver challenges, including caregiver burden, unmet needs, and system failures (breakdowns in caregiving support systems).^6,7^ Although these domains are often studied separately, they are likely to be closely interconnected in real-world caregiving experiences. For example, inadequate access to respite services or behavioral management resources may increase caregiver burden while simultaneously creating unmet support needs. Nevertheless, how these challenges co-occur and interact within individual caregiver experiences is understudied.^8–10^ Understanding these relationships can provide important insight into whether burden, unmet needs, and system failures function as separate problems or as parts of one connected caregiving challenge ecosystem, and this insight can inform more effective and targeted caregiver support.

Caregiver challenges are also not uniform. They vary by caregiver role, whether someone is a primary or secondary caregiver, and by relationship to the care recipient. Prior work has shown that primary caregivers often report higher levels of subjective burden than secondary caregivers, reflecting their greater involvement in day-to-day care activities. However, secondary caregivers may also experience clinically significant distress and caregiving-related challenges that are frequently overlooked in both research and support programs.^11–13^ Spouses and adult children also experience caregiving differently. Adult child caregivers report greater burden and lower quality of life, while spousal caregivers experience greater grief as the disease advances.^14^ Prior literature suggests substantial heterogeneity in caregiver experiences. Yet most interventions do not account for these differences.^15–17^

Most prior studies characterizing dementia caregiver challenges have relied on structured surveys, interviews, and small-sample qualitative methods such as thematic analysis.^15,16,18^ While these approaches have generated important insights, they are limited by small sample sizes, predefined categories, and social desirability bias. Online caregiver forums offer a complementary data source. Posts are written voluntarily and anonymously, capturing spontaneous expressions that may not surface in clinical settings or structured research. ALZConnected, a moderated forum operated by the Alzheimer’s Association, has been used in 28 published studies, demonstrating the value of this platform as a source of caregiver-generated narratives^7^

In this study, we used an LLM-based classification framework to analyze posts from three ALZConnected caregiver forums (general caregivers, spouse/partner caregivers, and caregivers of parents), using categories developed inductively from caregivers’ own language and refined against prior literature. Using this approach, we characterized the prevalence, distribution, and co-occurrence structure of caregiver burden, unmet needs, and system failures in online ADRD caregiving discussions. We hypothesized that these challenges would be commonly expressed among ADRD caregivers and that their prevalence and distribution would vary across caregiver roles and relationships to the care recipient. We further hypothesized that burden, unmet needs, and system failures would not occur independently but would form interconnected caregiver challenge ecosystems, with consistent patterns of co-occurrence linking challenges within and across domains. Specifically, we examined the prevalence and distribution of each domain, assessed differences across caregiver roles and relationship types, and mapped how challenges interconnect within and across domains using co-occurrence network analysis. By showing that burden, unmet needs, and system failures form interconnected challenge ecosystems that take different shapes across caregiver roles and relationships, this work informs the design of caregiver support interventions tailored to specific caregiving contexts and offers an analytic framework that can be applied to other caregiver text data.

## 2. Methods

### 2.1 Data Source and Collection

Data were collected from ALZConnected (https://alzconnected.org), an online peer support community operated by the Alzheimer’s Association for individuals caring for persons living with ADRD. Publicly available posts were extracted from three caregiver forums representing general caregivers, spouse/partner caregivers, and caregivers of parents, using an automated web-scraping pipeline. Structured data, including post titles, content, author identifiers, posting dates, and discussion tags, were extracted from each forum page. Full data collection procedures are described in our prior study^19^.

### 2.2 Data Processing

Posts were chronologically ranked and restricted to the most recent 3,000 entries per forum as of February 2, 2026. Caregiver-specific abbreviations and ADRD terminology were expanded into standardized clinical language. A hybrid de-identification framework combining named entity recognition and rule-based masking was applied to remove identifying information and dementia-related diagnoses or medication mentions. Posts classified as non-caregiver or former caregiver narratives were excluded, and only posts from active caregivers (primary or secondary) were retained for subsequent analyses. Full data processing procedures are described in our prior study.^19^

### 2.3 LLM-Based Classification

Claude 3.5 Sonnet was used for all content classification with deterministic inference settings (temperature = 0.0) to improve reproducibility via Stanford’s secure API environment.^20^ Classification focused on three domains: caregiver burden, unmet needs, and system failures (breakdowns in caregiving support systems).

Classification was performed in two sequential steps for each domain. Posts were first classified according to whether the domain was expressed (yes/no). Posts classified as expressing the domain were then assigned up to three predefined categories ranked by relevance. Because expression and category assignment were evaluated separately, some posts were classified as expressing a domain without receiving a specific category assignment. Prompts are provided in Method S1.

#### 2.3.1 Category Development

To avoid predefined category bias, initial prompts requested free-response descriptions of burden, unmet needs, and system failures, with up to three responses per post ranked by relevance. A random sample of 2,000 free-text responses per domain was then analyzed using Claude 3.5 Sonnet to inductively generate categories through thematic clustering. The resulting data-driven categories were reviewed against the existing literature to ensure comprehensive coverage, and categories not represented in the data but identified through literature review were added. Full category definitions are provided in Table S1. This inductive-deductive approach ensured that categories were grounded in caregivers’ own expressions while maintaining alignment with established literature.

The final classification schema comprised 9 burden categories, 12 unmet need categories, and 10 system failure categories. For burden, these covered physical exhaustion, emotional and psychological distress, decision-making and ethical burden, social isolation and relationship strain, financial and employment impact, administrative and care coordination burden, behavioral management and safety, loss of personal identity and freedom, and unequal family responsibility.^21–27^ For unmet needs, categories spanned emotional and psychological support, respite and caregiver support services, practical caregiving tasks and equipment, clinical care information and symptom management, healthcare navigation and care coordination, safety and behavior management, financial assistance and benefits access, legal authority and advance planning, family roles and conflict guidance, work and employment support, care transitions and placement, and end-of-life and grief support.^6,21,25,28–30^ System failure categories included financing and coverage failures, care coordination and provider communication failures, caregiver support service gaps, long-term care facility quality and access issues, legal and protective system failures, home care service quality and reliability, healthcare access and geographic barriers, emergency and crisis response inadequacies, workplace and employment support failures, and technology and infrastructure barriers.^28,31–41^ Posts were allowed up to three categories per domain to reflect the reality that caregivers often experience multiple concurrent challenges. Posts that did not fit any category were coded as other. Detailed prompts are provided in Method S1.

#### 2.3.2 Inter-model reproducibility

Because caregiver forum posts express inherently subjective experiences, there is no single ground truth against which LLM annotations can be validated.^42,43^ This study uses LLM-based classification as an exploratory, large-scale content analysis tool. To assess reproducibility, we compared Claude 3.5 Sonnet outputs with independently generated GPT-4o outputs on a randomly selected 10% of posts, using identical prompts and inference settings. Agreement was evaluated using Cohen’s kappa and exact match rates for categorical variables. Inter-model agreement serves as an indicator of classification stability across models rather than a validation of classification accuracy.

### 2.4 Analysis

For burden, unmet need, and system failure separately, we summarized overall expression, the distribution of coded categories, and the number of categories assigned per post (0-3). Category prevalence was examined overall and stratified by caregiver role (primary vs. secondary) and relationship to the care recipient. Relationship analyses were conducted at granular and broad levels (spousal vs. child). Heatmaps were used to summarize category-specific patterns across caregiver role and relationship type.

Overall expression was compared between primary and secondary caregivers and between spousal and child caregivers using Pearson χ² tests. Category-specific prevalence was compared using the same approach for each category, with Holm correction applied.

Co-occurrence among categories was examined using post-level network graphs, treating each pair of categories as connected if they were significantly associated at the post level. Nodes represented categories, and node size reflected prevalence. Pairwise associations were tested using Fisher’s exact tests, with Holm correction applied separately within each network block (within burden, within need, within failure, and across domains). Only pairs with Holm-adjusted P < 0.05 and |φ| ≥ 0.05 were displayed. We required both statistical significance after multiple-comparison correction and a minimum effect size threshold, so that networks reflect associations that are both reliable and non-trivial in magnitude rather than statistically significant but negligibly small. Within-domain networks summarized joint co-occurrence (percentage of posts mentioning both categories). An integrated network summarized significant cross-domain links among burden, need, and system-failure categories. All tests were two-sided. Significance was set at α = 0.05.

### 2.5 Ethics Statement

This study was approved by the Stanford University Institutional Review Board (IRB Protocol #83818). Informed consent and data use authorization were waived given the use of publicly available, de-identified online forum data. The study was classified as minimal risk under expedited review.

## 3. Results

### 3.1 Inter-model Reproducibility

Inter-model reproducibility between Claude 3.5 Sonnet and GPT-4o was assessed on a random 10% of posts using identical prompts and inference settings. Binary classification of burden expression showed high consistency (κ=0.81, exact match=94.0%), as did unmet needs (κ=0.79, exact match=94.5%) and system failure detection (κ=0.77, exact match=90.0%).

### 3.2 Sample Characteristics

A total of 7,198 posts from primary and secondary caregivers were analyzed, including primary caregivers (n=4515, 62.7%) and secondary caregivers (n=2683, 37.3%). Among posts with identifiable relationship categories, child caregivers were the largest group (n=3418), followed by spousal caregivers (n=2405) and other caregivers (n=523). This sample is drawn from the same source dataset described in our prior study^19^, which reports additional cohort characteristics including caregiving objectives, emotional valence, and posting behavior.

### 3.3 Burden

Descriptive patterns of caregiver burden expression are shown in Figure 1. Burden was expressed in 89.0% of posts (6409/7198; Figure 1A). Multiple burden categories were common: 34.2%, 30.9%, and 23.9% of posts were assigned three, two, or one burden code, respectively (Figure 1B). The most frequent categories were managing difficult behaviors or safety risks (24.4%), emotional and psychological distress (21.3%), and decision-making and ethical burden (20.4%); physical exhaustion was least common (2.3%; Figure 1E).

**Figure 1.**
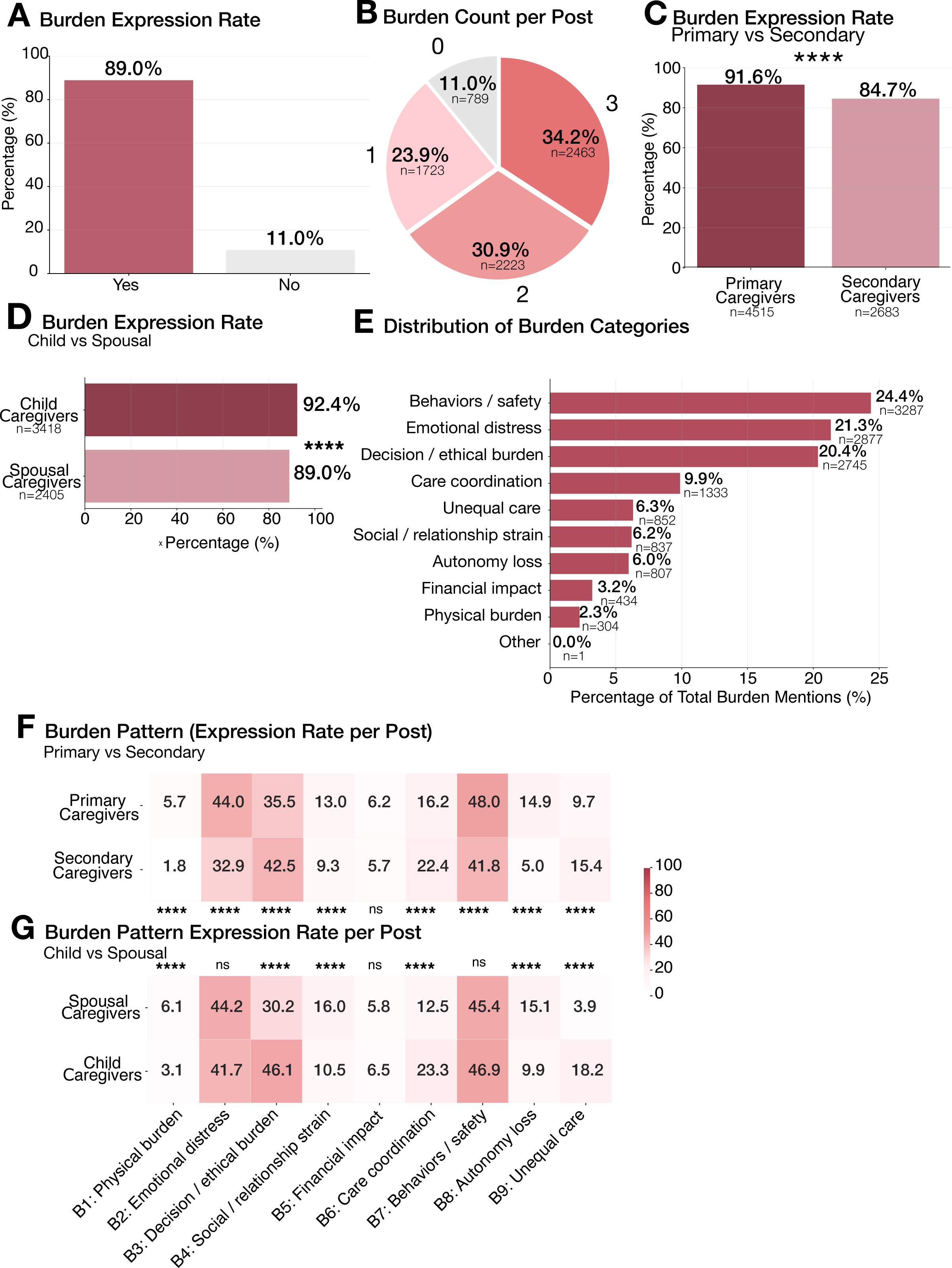
Prevalence, distribution, and subgroup patterns of caregiver burden among ADRD caregivers. (A) Overall burden expression rate. (B) Number of burden categories assigned per post among posts expressing burden. (C) Burden expression rates among primary and secondary caregivers. (D) Burden expression rates among child and spousal caregivers. (E) Distribution of burden categories. (F) Burden category expression rates among primary and secondary caregivers. (G) Burden category expression rates among child and spousal caregivers. Heatmaps display the percentage of posts within each subgroup expressing each burden category. Full category definitions are provided in Method S1. ****P < 0.0001.

Primary caregivers expressed burden more often than secondary caregivers (91.6% vs. 84.7%; P<0.0001; Figure 1C). Burden category profiles also differed by caregiver role. Eight of nine categories showed significant group differences (P<0.0001; Figure 1F). Primary caregivers reported higher rates of physical burden, emotional distress, social and relationship strain, behavior/safety management, and autonomy loss. Secondary caregivers reported higher rates of decision-making and ethical burden, care coordination, and unequal care distribution.

Child caregivers expressed burden more often than spousal caregivers (92.4% vs. 89.0%; P<0.0001; Figure 1D). Category profiles also differed by broad relationship type (six of nine categories; P<0.0001; Figure 1G). Spousal caregivers reported higher rates of physical burden, social and relationship strain, and autonomy loss. Child caregivers reported higher rates of decision-making and ethical burden, care coordination, and unequal care. Granular relationship-specific patterns are shown in Figures S1 and S2.

### 3.4 Unmet Needs

Descriptive patterns of caregiver needs expression are shown in Figure 2. Needs were expressed in 93.3% of posts (6714/7198; Figure 2A). Multiple need categories were common: 56.8%, 29.6%, and 6.9% of posts were assigned three, two, or one need code, respectively (Figure 2B). The most frequent need categories were clinical information (20.0%), emotional support (16.8%), and safety management (14.9%), whereas work support (0.7%) and end-of-life support (3.0%) were least common (Figure 2E).

**Figure 2.**
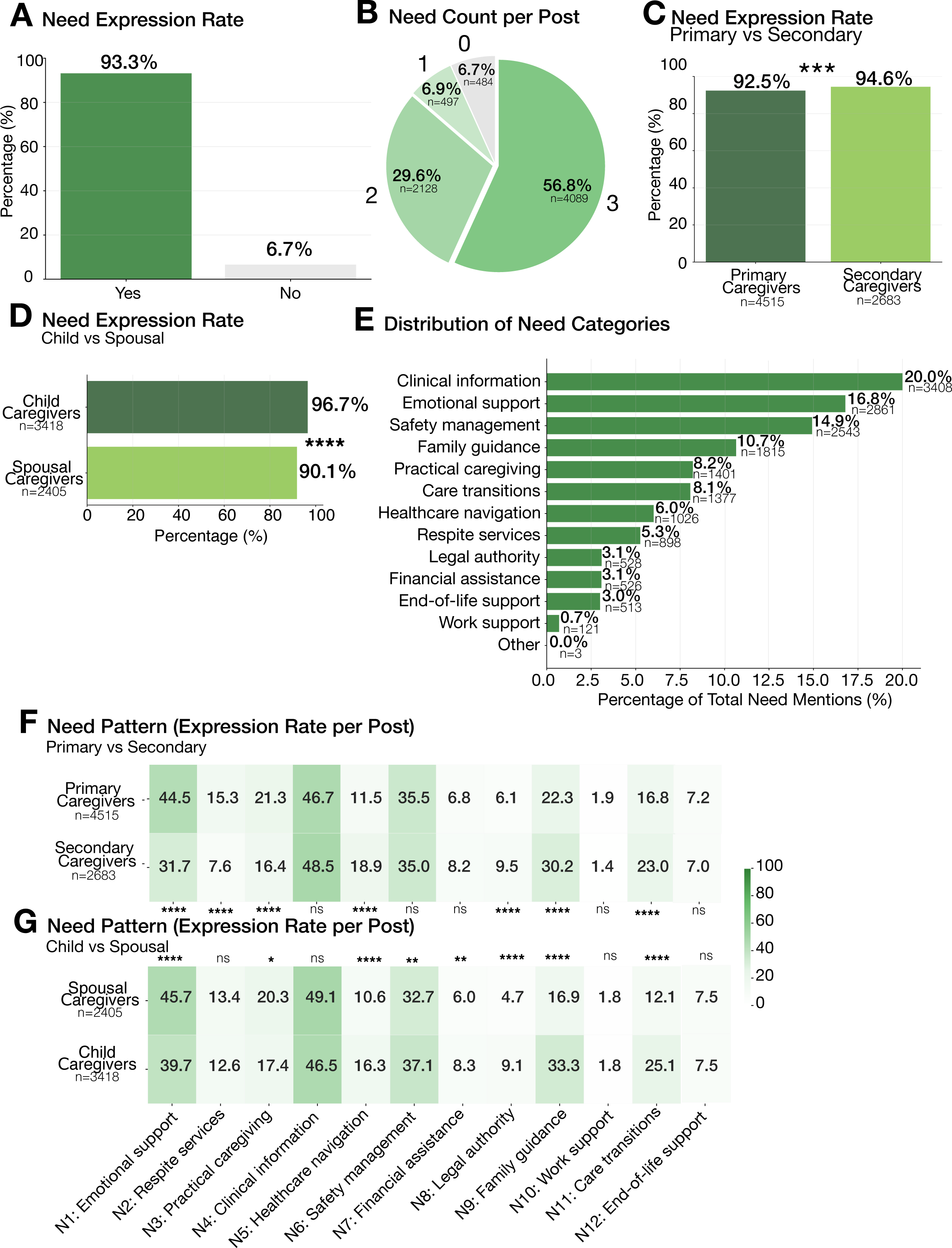
Prevalence, distribution, and subgroup patterns of unmet needs among ADRD caregivers. (A) Overall unmet need expression rate. (B) Number of unmet need categories assigned per post among posts expressing unmet needs. (C) Unmet need expression rates among primary and secondary caregivers. (D) Unmet need expression rates among child and spousal caregivers. (E) Distribution of unmet need categories. (F) Unmet need category expression rates among primary and secondary caregivers. (G) Unmet need category expression rates among child and spousal caregivers. Heatmaps display the percentage of posts within each subgroup expressing each unmet need category. Full category definitions are provided in Method S1. Statistical significance for category-specific group comparisons was assessed using Pearson χ² tests with Holm correction for multiple comparisons. ***P < 0.001; ****P < 0.0001.

Primary caregivers expressed needs less often than secondary caregivers (92.5% vs. 94.6%; P<0.0001; Figure 2C). Need category profiles also differed by caregiver role. Eight of twelve categories showed significant group differences (Figure 2F). Compared with secondary caregivers, primary caregivers more frequently expressed needs related to emotional support, respite services, practical caregiving, and family guidance. Secondary caregivers more frequently expressed needs related to healthcare navigation, safety management, legal authority, and care transitions. Clinical information, financial assistance, work support, and end-of-life support needs were comparable between groups.

Child caregivers expressed needs more often than spousal caregivers (96.7% vs. 90.1%; P<0.0001; Figure 2D). Need category profiles also differed by broad relationship type, with eight of twelve categories showing significant differences (Figure 2G). Spousal caregivers more frequently expressed needs related to emotional support, clinical information, and practical caregiving. In contrast, child caregivers more frequently expressed needs related to healthcare navigation, safety management, financial assistance, legal authority, family guidance, and care transitions. Granular relationship-specific patterns are shown in Figures S3 and S4.

### 3.5 System Failures

Descriptive patterns of system failure expression are shown in Figure 3. System failures were described in 34.8% of posts (2505/7198; Figure 3A). Multiple system failure categories were common: 20.5%, 8.2%, and 3.8% of posts were assigned three, two, or one system failure code, respectively (Figure 3B). The most frequently described system failure categories were caregiver support gaps (33.0%), care coordination failures (17.3%), long-term care facility issues (12.8%), and emergency or crisis-related failures (12.7%), whereas technology and infrastructure failures were least common (0.8%; Figure 3E).

**Figure 3.**
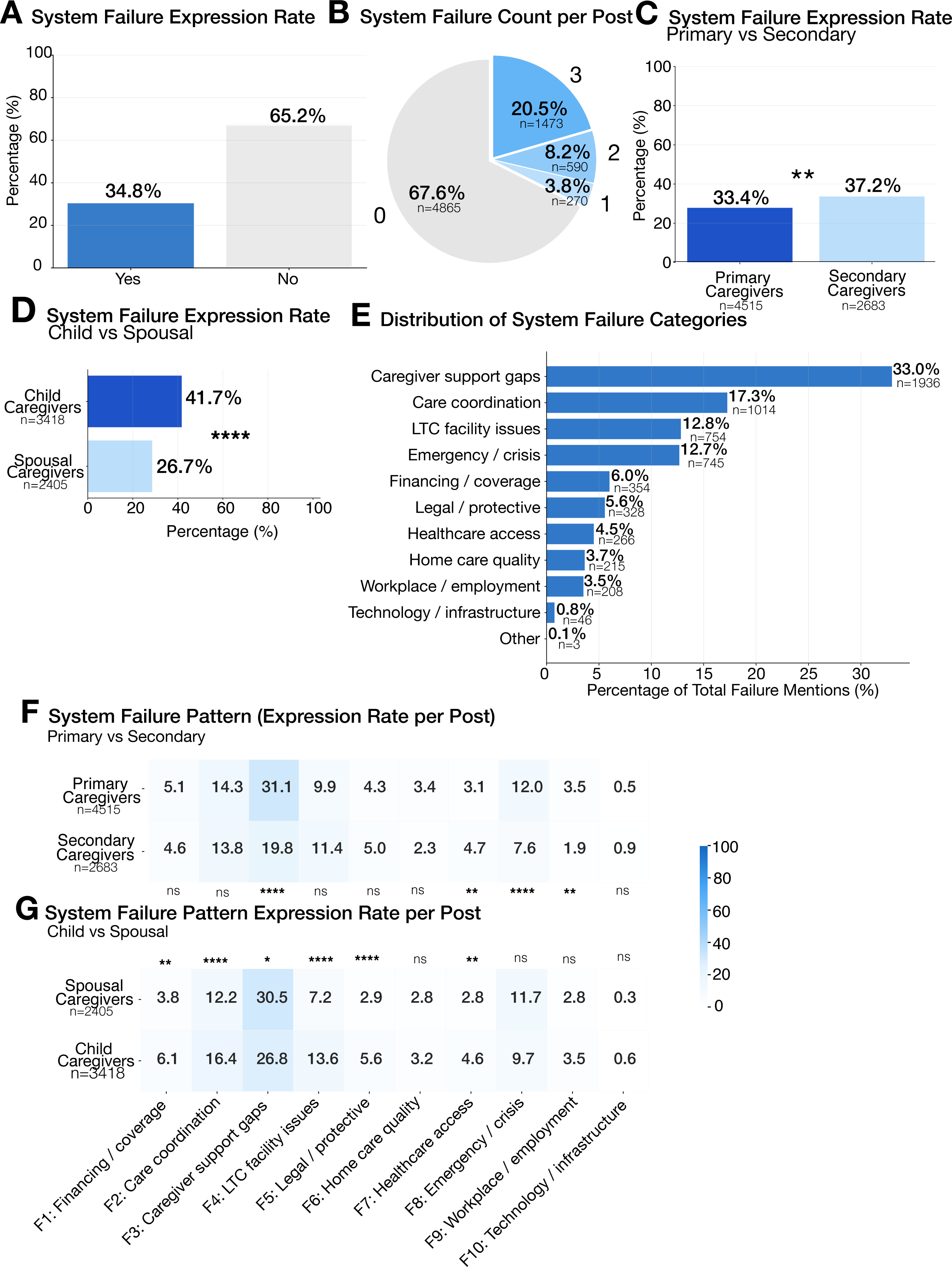
Prevalence, distribution, and subgroup patterns of system failures among ADRD caregivers. (A) Overall system failure expression rate. (B) Number of system failure categories assigned per post among posts expressing system failures. (C) System failure expression rates among primary and secondary caregivers. (D) System failure expression rates among child and spousal caregivers. (E) Distribution of system failure categories. (F) System failure category expression rates among primary and secondary caregivers. (G) System failure category expression rates among child and spousal caregivers. Heatmaps display the percentage of posts within each subgroup expressing each system failure category. Full category definitions are provided in Method S1. Statistical significance for category-specific group comparisons was assessed using Pearson χ² tests with Holm correction for multiple comparisons. **P < 0.01; ****P < 0.0001.

Secondary caregivers described system failures more often than primary caregivers (37.2% vs. 33.4%; P<0.01; Figure 3C). System failure category profiles also differed by caregiver role. Four of ten categories showed significant group differences (Figure 3F). Compared with secondary caregivers, primary caregivers more frequently described caregiver support gaps, emergency or crisis-related failures, and workplace or employment-related challenges. In contrast, secondary caregivers more frequently described healthcare access barriers. No significant differences were observed for financing and coverage, care coordination, long-term care facility issues, legal or protective failures, home care quality, or technology and infrastructure failures.

Child caregivers described system failures substantially more often than spousal caregivers (41.7% vs. 26.7%; P<0.0001; Figure 3D). System failure category profiles also differed by broad relationship type, with six of ten categories showing significant differences (Figure 3G). Compared with spousal caregivers, child caregivers more frequently described financing and coverage problems, care coordination failures, long-term care facility issues, legal or protective failures, and healthcare access barriers. In contrast, spousal caregivers more frequently described caregiver support gaps. Granular relationship-specific patterns are shown in Figures S5 and S6.

### 3.6 Interconnected Caregiver Challenges

Having established that burden, unmet needs, and system failures each vary systematically by caregiver role and relationship, we next tested whether these domains occur independently or form part of one connected system of caregiving challenges. Co-occurrence analyses revealed systematic clustering of burden, need, and system-failure categories, indicating that these are not separate problems but interconnected caregiver challenge ecosystems (Figure 4). Networks displayed only category pairs with significant post-level co-occurrence (Holm-adjusted P < 0.05; |φ| ≥ 0.05).

**Figure 4.**
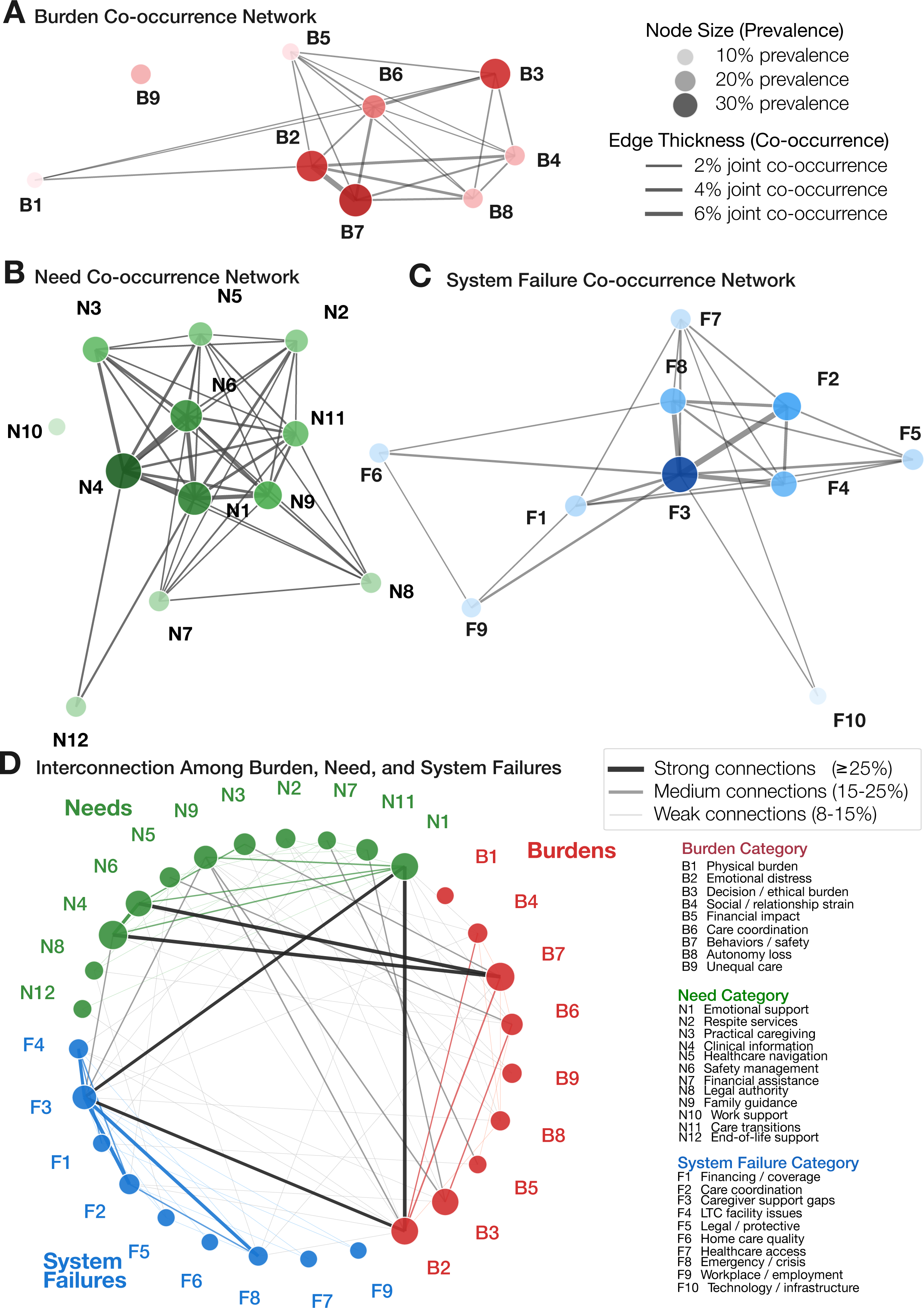
Co-occurrence networks of caregiver burden, unmet needs, and system failures among ADRD caregivers. (A) Co-occurrence network of caregiver burden categories. (B) Co-occurrence network of unmet need categories. (C) Co-occurrence network of system failure categories. (D) Integrated cross-domain network showing connections among burden, unmet need, and system failure categories. Nodes represent individual categories, with node size proportional to category prevalence. Edges represent statistically significant pairwise co-occurrence relationships identified using Fisher’s exact tests with Holm correction. In panels A-C, edge thickness corresponds to the proportion of posts in which both categories were co-expressed. In panel D, edge thickness represents the strength of association, categorized as weak (8%-15%), medium (15%-25%), or strong (≥25%) co-occurrence. Only categories with a prevalence of at least 2.5% were included in the network visualization. Full category definitions are provided in Method S1.

The burden co-occurrence network included 9 categories connected by 22 significant edges (22 of 36 pairwise associations; network density = 0.61), indicating substantial overlap among burden experiences (Figure 4A). Emotional distress, behavior/safety management, decision-making/ethical burden, and care coordination were among the most interconnected burden categories.

The need network included 12 categories connected by 43 significant edges (43 of 66 pairwise associations; density = 0.65), with emotional support, clinical information, safety management, healthcare navigation, and family guidance forming a densely connected core of co-occurring support needs (Figure 4B).

The system-failure network showed the highest within-domain interconnectedness, with 10 categories connected by 28 significant edges (28 of 45 pairwise associations; density = 0.62; Figure 4C). Caregiver support gaps and care coordination failures were the most interconnected failure categories.

The integrated network demonstrated extensive cross-domain linkage among 31 burden, need, and system-failure categories (84 significant connections; Figure 4D), directly supporting the ecosystem framing: challenges in one domain are systematically tied to challenges in the others rather than occurring on separate tracks. The strongest cross-domain co-occurrences linked behavior/safety burden with safety-management need (21.7% of posts) and emotional burden with emotional-support need (20.9%). Additional strong cross-domain links connected emotional distress and decision-making/ethical burden with emotional support, healthcare navigation, caregiver support gaps, and care coordination failures.

## 4. Discussion

This study characterized the prevalence, distribution, and co-occurrence structure of caregiver burden, unmet needs, and system failures in a large sample of ADRD caregiver forum posts. Burden and unmet needs were expressed in the vast majority of posts, while system failures were described in more than one-third of posts. The prevalence and distribution of these challenges varied systematically across caregiver roles and relationships to the care recipient, highlighting the heterogeneous nature of the caregiving experience. Importantly, burden, unmet needs, and system failures did not emerge as independent dimensions of caregiving. Instead, they formed interconnected caregiver challenge ecosystems, systematic patterns of co-occurrence linking emotional, practical, and systemic difficulties within individual caregivers’ experiences. These findings underscore the need for support strategies that account for caregiver context and for the interconnected nature of caregiver challenges, rather than addressing burden, needs, and system failures as separate problems.

The co-occurrence networks provide additional insight into the structure of caregiver experiences. We found that burden, unmet needs, and system failures frequently co-occurred both within and across domains. Prior studies have largely examined these challenges in isolation.^44,45^ Our findings suggest instead that caregivers commonly experience and describe these domains together. These clusters also suggest potential explanations of why the domains link together, though these are interpretations of the pattern rather than causal claims. The strongest cross-domain connection, between behavioral and safety burden and safety-management needs, may reflect a documented gap in dementia care. Caregivers managing behavioral symptoms often express difficulty handling them and limited understanding of how to access relevant support services.^46,47^ The co-occurence of emotional distress and decision-making burden with caregiver support gaps and care coordination failures is similarly consistent with prior work describing how emotionally difficult caregiving decisions frequently coincide with systemic breakdowns, including fragmented care and coordination gaps between providers.^30,48^ These findings suggest that burden, unmet needs, and system failures are not three separate problems, but parts of one connected system: a challenge in one domain tends to appear alongside challenges in the others. This may help explain why caregiver distress remains high despite decades of caregiver support research. Interventions addressing a single domain in isolation may leave connected challenges in other domains unaddressed.

The most commonly expressed burden categories were behavioral and safety burden, emotional distress, and decision-making and ethical burden. This pattern is consistent with prior studies identifying behavioral and psychological symptoms of dementia as major drivers of caregiver distress.^26,49,50^ The prominence of decision-making and ethical burden further highlights the complex choices caregivers frequently face regarding autonomy, safety, medical treatment, and long-term care. Beyond burden, caregivers most frequently expressed unmet needs related to clinical information, emotional support, and safety and behavior management. These findings align with previous work showing that caregivers often feel inadequately prepared to manage disease progression, behavioral symptoms, and complex care decisions and frequently report unmet needs for information and emotional support.^6,51^ Notably, caregiver support service gaps emerged as the most commonly described system failure. While prior studies have documented persistent gaps in caregiver resources and support programs, our findings suggest that challenges related to caregiver support remain a central feature of caregiver experiences despite growing recognition of the importance of caregiver-focused services and care navigation programs.^30,48,52^

Primary and secondary caregivers showed distinct challenge profiles across domains. Primary caregivers expressed burden more frequently, particularly emotional distress and loss of autonomy, consistent with prior studies documenting high levels of subjective burden, anxiety, depression, and role strain among individuals providing the majority of hands-on care.^11,53^ In contrast, secondary caregivers expressed more unmet needs and system failures, healthcare navigation needs, family conflict guidance, and care coordination challenges. One possible explanation is that secondary caregivers are often involved in coordinating care, communicating with family members, and interacting with healthcare and long-term care systems rather than providing day-to-day care. This interpretation is consistent with evidence that dementia care frequently requires coordination across multiple providers, services, and support systems, creating substantial logistical and informational challenges for caregivers.^30,40,48^ These findings suggest that secondary caregivers may be more involved in navigating healthcare and long-term care systems and coordinating aspects of care, whereas primary caregivers experience greater direct emotional and practical burden.

Differences across relationship types further highlight the heterogeneous nature of ADRD caregiving. Child caregivers expressed the highest levels of burden, unmet needs, and system failures across broad relationship groups. This finding is consistent with prior studies showing that adult child caregivers often report greater burden and lower quality of life while balancing caregiving responsibilities with employment, childcare, and other family obligations.^14,54–56^ Our findings extend this literature by suggesting that these differences may reflect not only greater caregiving demands but also distinct caregiving roles. Compared with spousal caregivers, child caregivers more frequently described decision-making burden, healthcare navigation needs, care coordination challenges, legal authority concerns, and system failures. These patterns suggest that child caregivers are often required to navigate complex healthcare and long-term care systems while coordinating care across multiple stakeholders. In contrast, spousal caregivers more frequently expressed challenges related to direct caregiving and emotional support. Together, these findings indicate that caregiver experiences differ not only in magnitude but also in the types of challenges encountered, underscoring the need for relationship-specific approaches to caregiver support.

This study has several strengths. It uses naturalistic, unfiltered caregiver narratives from a dementia-specific online community, capturing experiences that may not be fully represented in structured surveys. Large language model-based classification allowed us to code a large volume of caregiver narratives at a level of context sensitivity that would be difficult to achieve with rule-based or purely manual coding. Categories were developed through an inductive process grounded in both the data and existing literature, and the resulting coding scheme and network analysis approach could reasonably be applied to other caregiver text data beyond this dataset. The co-occurrence network approach itself offers a way to visualize and quantify the interconnectedness of caregiver challenges, moving beyond prevalence statistics to characterize the structural relationships among domains, which is the central contribution of this work.

These findings have several practical implications. Because burden, unmet needs, and system failures tend to arise together rather than separately, caregiver assessments that screen for each domain as a separate checklist item may miss these intricate relationships. An integrated assessment could instead flag likely co-occurring challenges once one domain is identified, for example, prompting a follow-up on safety-management needs and caregiver support availability when a caregiver reports difficulty managing behavioral symptoms. Similarly, AI-enabled care navigation tools could use these documented co-occurrence patterns to anticipate related challenges rather than responding only to what a caregiver has explicitly reported, potentially surfacing relevant resources earlier in the caregiving trajectory. Because the shape of these ecosystems differed by caregiver role and relationship, such tools would likely need to be tailored accordingly rather than applying a single model across all caregivers.

Several limitations should be noted. First, the sample reflects caregivers who are able and willing to participate in an online support community and therefore is not representative of all ADRD caregivers. Individuals are likely to post when seeking advice, emotional support, or assistance during periods of increased caregiving challenges.^57,58^ Consequently, the prevalence of burden, unmet needs, and system failures reported here should not be interpreted as population-level estimates. In addition, online forum users likely skew younger and more digitally literate, so older spousal caregivers and individuals with limited digital access may be underrepresented in this sample.^59,60^ Second, all data were obtained from a single online platform, and discourse patterns may differ across caregiver communities.^57^ Third, relationship categories and caregiving contexts were inferred from self-reported narratives rather than verified records. Fourth, although inter-model agreement was high, LLM-based classification remains subject to prompt sensitivity and model-specific error, particularly for ambiguous posts, and agreement between models indicates reproducibility rather than validated accuracy. Finally, the cross-sectional nature of the data limits inference regarding how caregiver experiences evolve over time.

In conclusion, this large-scale analysis of dementia caregiver forum posts reveals that burden and unmet needs are near-universal, that system failures are prevalent but distinct in their distribution and triggers, and that all three domains form interconnected caregiver challenge ecosystems rather than independent problems. Behavioral management, emotional distress, and decision-making burden form the core burden experience specific to dementia caregiving, while caregiver support service gaps represent the most broadly connected systemic failure. The systematic heterogeneity across primary and secondary caregivers and across relationship types provides an empirical basis for moving beyond one-size-fits-all approaches toward role-specific, ecosystem-aware caregiver support.

**Table 1.** Prevalence of caregiver burden, unmet needs, and system failure expression by caregiver role and relationship.

| Category | n | Burden expressed, n (%) | Unmet need expressed, n (%) | System failure expressed, n (%) |
| --- | --- | --- | --- | --- |
| **Overall** | 7,198 | 6,409 (89.0) | 6,714 (93.3) | 2,505 (34.8) |
| **Caregiver role** |  |  |  |  |
| Primary caregiver | 4,515 | 4,137 (91.6) | 4,177 (92.5) | 1,507 (33.4) |
| Secondary caregiver | 2,683 | 2,272 (84.7) | 2,537 (94.6) | 998 (37.2) |
| **Relationship (broad)** |  |  |  |  |
| Spousal | 2,405 | 2,140 (89.0) | 2,166 (90.1) | 643 (26.7) |
| Child | 3,418 | 3,159 (92.4) | 3,305 (96.7) | 1,427 (41.7) |
| Other | 523 | 452 (86.4) | 493 (94.3) | 217 (41.5) |
| **Relationship (granular)** |  |  |  |  |
| Spouse/partner : Husband | 1,731 | 1,560 (90.1) | 1,580 (91.3) | 510 (29.5) |
| Spouse/partner : Wife | 654 | 566 (86.5) | 567 (86.7) | 126 (19.3) |
| Child caregiver: Mother | 2,631 | 2,436 (92.6) | 2,539 (96.5) | 1,115 (42.4) |
| Child caregiver: Father | 778 | 719 (92.4) | 759 (97.6) | 309 (39.8) |
| Sibling | 113 | 93 (82.3) | 102 (90.3) | 49 (43.4) |
| In-law | 105 | 92 (87.6) | 95 (90.5) | 42 (40.0) |
| In-law: Mother-in-law | 75 | 68 (90.7) | 73 (97.3) | 26 (34.7) |
| In-law: Father-in-law | 22 | 19 (86.4) | 22 (100.0) | NA |
| Other relative | 71 | 63 (88.7) | 68 (95.8) | 36 (50.7) |
| Grandchild caregiver: Grandmother | 62 | 56 (90.3) | 59 (95.2) | 24 (38.7) |
| Grandchild caregiver: Grandfather | 23 | 22 (95.7) | 23 (100.0) | 11 (47.8) |
| Professional caregiver | 31 | 23 (74.2) | 30 (96.8) | 11 (35.5) |
| Friend | 21 | 16 (76.2) | 21 (100.0) | NA |

## 7. Data Availability Statement

The underlying posts are publicly available on ALZConnected (https://alzconnected.org). The processed dataset used in this analysis is not publicly available. Researchers interested in the data may contact the corresponding author.

