## Supplementary Material for "Interconnected Challenges in Dementia Caregiving: A Co-occurrence Network Analysis of Burden, Unmet Needs, and System Failures Among Caregivers"

**Method S1. LLM Classification Prompts**

Classification was conducted in three phases: (1) free-response detection across all three companion papers (2) inductive thematic clustering to generate categories and (3) category-level classification using the finalized schema. Phase 1 classification was run jointly with other prompts used for companion papers [citation]. Prompts specific to Paper 1 and Paper 3 are reported in their respective manuscripts. Phase 1 prompts were used to detect the presence of each domain and to generate free-response descriptions for category development. Phase 2 prompts were used for category development only. Phase 3 prompts were used for category-level classification in all analyses reported in this study.

*Phase 1: Free-Response Classification*

*Prompt 1A: Caregiver Burden and Unmet Needs*

```
### Analyze below post for caregiver burden and needs following below instructions

1. Caregiver Burden
– Decide if the author expresses any burden.
 - If yes, set "burden_expressed" to 1 and list up to 3 needs in the order of priority with a short explanation. If you cannot determine the burden, mention NA.
 - If no, set "burden_expressed" to 0 and set "burden_details" NA.

2. Unmet Needs
– Decide if the author states any unmet needs.
 - If yes, set "needs_expressed" to 1 and list up to 3 needs in the order of priority with a short explanation. If you cannot determine the need, mention NA.
 - If no, set "needs_expressed" to 0 and set "needs_details" NA.

IMPORTANT: Return only the valid JSON object. Use double quotes for all keys and string values. Base all analysis strictly on the content explicitly provided in the post. Do not infer information not stated in the text.

Example output JSON format:
{
"burden_and_needs": {
    "burden_expressed": int (0|1),
    "burden 1": "",
    "burden 2": "",
    "burden 3": "",
    "burden_justification": "",
    "need_expressed": int (0|1),
    "need 1": "",
    "need 2": "",
    "need 3": "",
    "needs_justification": ""
  }
}

### Post Content:
[POST TEXT]
```

*Prompt 1B: System failures*

```
 ### Identify any "System Failures" problems with healthcare, social services, insurance, etc following below instructions
    • If yes, set "system_failures.mentioned" to 1 and list up to 3 system failures in the order of priority. If you cannot determine the need, mention NA.
    • If no, set "system_failures.mentioned" to 0 and mention "NA" for list of "failures" and "justification".

    IMPORTANT: Return only the valid JSON object. Use double quotes for all keys and string values. Base all analysis strictly on the content explicitly provided in the post. Do not infer information not stated in the text.

    Example output JSON format:
    {

    "system_failures": {
        "mentioned": int (0|1),
        “failure 1”: “”,
        “failure 2”: “”,
        “failure  3”: “”,
        "justification": ""                       // brief reasoning
        }
    }

    ### Post Content:
 [POST TEXT]
```

*Phase 2: Inductive Thematic Clustering*

Initial thematic clustering was prompted using a minimal context-only prompt to allow inductive category generation without predefined structure. The resulting clusters were then reviewed and refined by the research team.

*Prompt 2A: Burden Clustering*

CAREGIVER BURDENS CATEGORIZATION ANALYSIS

CONTEXT: You are analyzing challenges, stresses, and difficulties experienced by caregivers from an online caregiver support forum. These represent real experiences from caregivers.

ANALYSIS FOCUS: Types of stress, strain, and challenges (physical, emotional, financial, social, etc.)

SAMPLE DATA:

[SAMPLE DATA]

*Prompt 2B: Unmet Needs Clustering*

CAREGIVER NEEDS CATEGORIZATION ANALYSIS

CONTEXT: You are analyzing support, resources, and assistance that caregivers require from an online caregiver support forum. These represent real experiences from caregivers.

ANALYSIS FOCUS: Types of support needed (informational, emotional, practical, financial, respite, etc.)

SAMPLE DATA:

[SAMPLE DATA]

*Prompt 2C: System Failures Clustering*

CAREGIVER FAILURES CATEGORIZATION ANALYSIS

CONTEXT: You are analyzing systemic, institutional, and structural breakdowns affecting caregiving from an online caregiver support forum. These represent real experiences from caregivers.

ANALYSIS FOCUS: System failures (healthcare, social services, legal, financial, family support, etc.)

SAMPLE DATA:

[SAMPLE DATA]

*Phase 3: Category Classification*

*Prompt 3A: Burden Clustering*

```
TASK: Classify the caregiver forum post into the burden category that best fits.

CATEGORIES AND DEFINITIONS:
1. Physical Health and Exhaustion Burden — Physical exhaustion, health deterioration, or injury resulting from caregiving tasks
2. Emotional and Psychological Distress Burden — Generalized depression, anxiety, grief, or emotional exhaustion from the caregiving role
3. Decision-Making and Ethical Burden — Guilt, moral conflict, or distress arising from specific care decisions, including placement, end-of-life choices, diagnosis disclosure, or therapeutic deception
4. Social Isolation and Relationship Strain Burden — Loss of social connections, strained relationships, or deterioration of the pre-illness relationship with the care recipient
5. Financial and Employment Impact Burden — Job loss, reduced income, out-of-pocket care costs, or career sacrifice
6. Administrative and Care Coordination Burden — Managing health systems, insurance, legal documents, scheduling, or cross-provider care coordination
7. Behavioral Management and Safety Burden — Managing difficult behaviors or safety risks including aggression, wandering, falls, delusions, or resistance to care
8. Loss of Personal Identity and Freedom Burden — Loss of autonomy, personal identity, or life goals resulting from structural confinement to the caregiving role
9. Unequal Family Responsibility and Lack of Support Burden — Disproportionate caregiving responsibility with insufficient support from family members
10. Other Burden — Burden not adequately represented by categories 1–9

INSTRUCTIONS:
Prefer one category. Assign up to three only if each burden is explicitly stated and equally central.
If multiple categories apply, order by prominence.
Use only the post text. Do not infer unstated facts.
Use 10 only if none of 1–9 fit.

IMPORTANT: Return only the valid JSON object. Use double quotes for all keys and string values.

Example output JSON format:
  "expressed_burden":
  {
       "burden_1": int (1|2|3|4|5|6|7|8|9|10)
       "burden_2": int (1|2|3|4|5|6|7|8|9|10)
       "burden_3": int (1|2|3|4|5|6|7|8|9|10)
  }

### Post to classify:
```

*Prompt 3B: Unmet Need Categories*

```
TASK: Classify the caregiver forum post into need category(ies) that best fit.

CATEGORIES:
1. Emotional and psychological support
2. Respite and caregiver support services
3. Practical caregiving tasks, equipment, and daily routines
4. Clinical care information, symptom management, and communication strategies
5. Healthcare navigation and care coordination
6. Safety and behavior management
7. Financial assistance and benefits access
8. Legal authority and advance planning
9. Family roles, conflict, and boundary setting
10. Work and employment support
11. Care transitions, placement, and housing
12. End-of-life, hospice, and grief support (includes anticipatory grief)
13. Other need

INSTRUCTIONS:
- If the post contains multiple needs from clearly different domains, assign up to 3 categories, ordered by prominence.
- Use only the post text. Do not infer unstated facts. Do not assign a category based on the care recipient's condition alone; focus on what the caregiver is asking for.
- Use 13 only if none of 1–12 fit.

IMPORTANT: Return only the valid JSON object. Use double quotes for all keys and string values.

Example output JSON format:
  "needs":
  {
    "need_1": int (1|2|3|4|5|6|7|8|9|10|11|12|13)
    "need_2": int (1|2|3|4|5|6|7|8|9|10|11|12|13)
    "need_3": int (1|2|3|4|5|6|7|8|9|10|11|12|13)
  }

### Post to classify:
[POST TEXT]
```

*Prompt 3C: System Failure Categories*

```
TASK: Classify the caregiver forum post into the systemic failure category that best fits.

CATEGORIES:
1. Financing/coverage failures: inability to afford or obtain coverage for needed care or supports
2. Care coordination/provider communication failures: poor handoffs, inconsistent information, or weak communication across providers or with families
3. Caregiver support service gaps: lack of services designed to support family caregivers
4. Long-term care facility quality/access issues: quality, safety, staffing, or access problems in residential care settings
5. Legal/protective system failures: gaps in legal protection, authority, or oversight for vulnerable adults
6. Home care service quality/reliability issues: unreliable or poor-quality in-home care services
7. Healthcare access/geographic barriers: limited local availability of services due to distance, provider shortages, or transportation
8. Emergency/crisis response inadequacies: inadequate system pathways for health or behavioral crises
9. Workplace/employment support failures: insufficient employer policies or legal protections for workers with caregiving duties
10. Technology/infrastructure barriers: digital or infrastructure barriers that impede care or communication
11. Other systemic failures: systemic failure present but does not fit 1–10

INSTRUCTIONS:
If the post contains multiple failures from clearly different domains, assign up to 3 categories, ordered by prominence.
Base classification on post text only. Do not infer unstated facts.
Use 11 only if 1–10 do not fit.

IMPORTANT: Return only the valid JSON object. Use double quotes for all keys and string values.

Example output JSON format:
  "systemic_failures":
  {
    "systemic_failure_1": int (1|2|3|4|5|6|7|8|9|10|11)
    "systemic_failure_2": int (1|2|3|4|5|6|7|8|9|10|11)
    "systemic_failure_3": int (1|2|3|4|5|6|7|8|9|10|11)
  }

### Post to classify:
[POST TEXT]
```

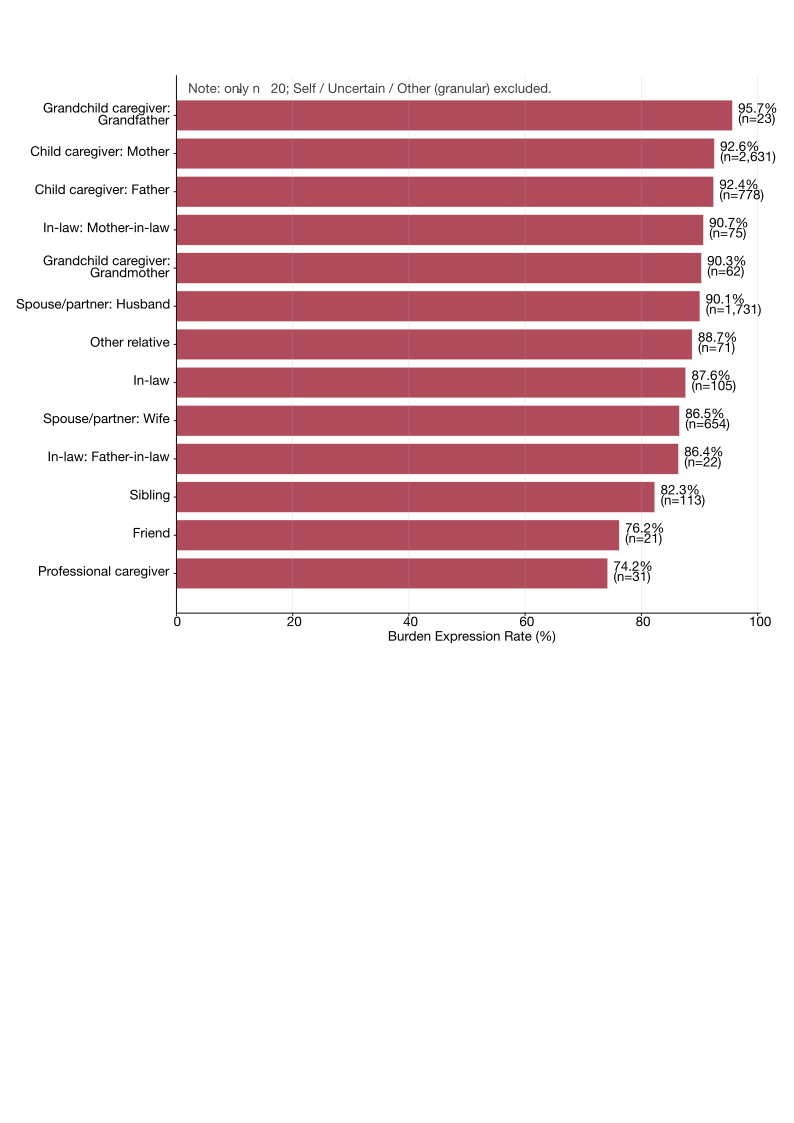

Figure S1. Overall burden expression across granular caregiver relationship categories.

Overall burden expression rates are shown for each caregiver relationship subgroup. Burden expression was defined as the presence of any caregiver burden. Only relationship categories with n ≥ 20 posts are displayed. Granular categories classified as self, uncertain, or other were excluded. Percentages indicate the proportion of posts within each relationship category expressing caregiver burden, and numbers in parentheses indicate the number of posts in each subgroup.

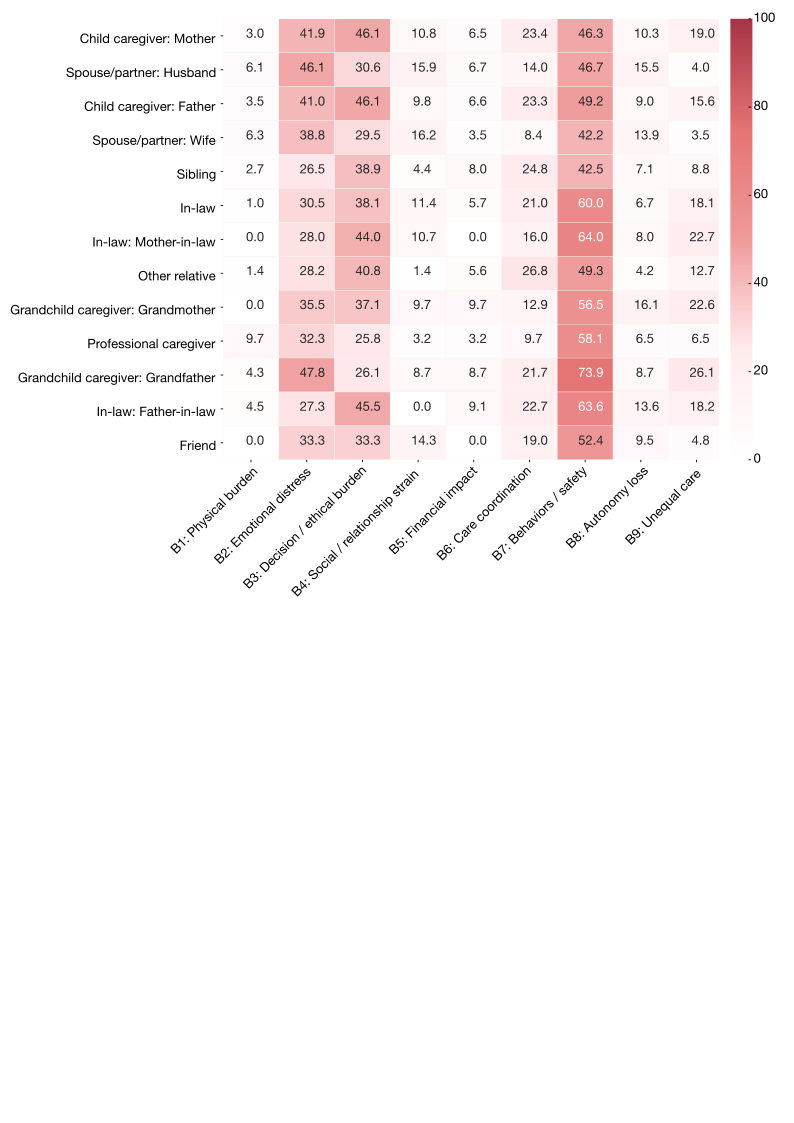

Figure S2. Burden category expression by granular caregiver relationship category.

Heatmap showing the percentage of posts within each caregiver relationship subgroup expressing each burden category. Percentages are calculated among posts within each relationship subgroup. Darker shading indicates a higher proportion of posts expressing the corresponding burden category. Only relationship categories with n ≥ 20 posts are shown. Granular categories classified as self, uncertain, or other were excluded. Burden categories include physical burden (B1), emotional distress (B2), decision-making and ethical burden (B3), social and relationship strain (B4), financial impact (B5), care coordination (B6), behavior and safety management (B7), autonomy loss (B8), and unequal care responsibility (B9).

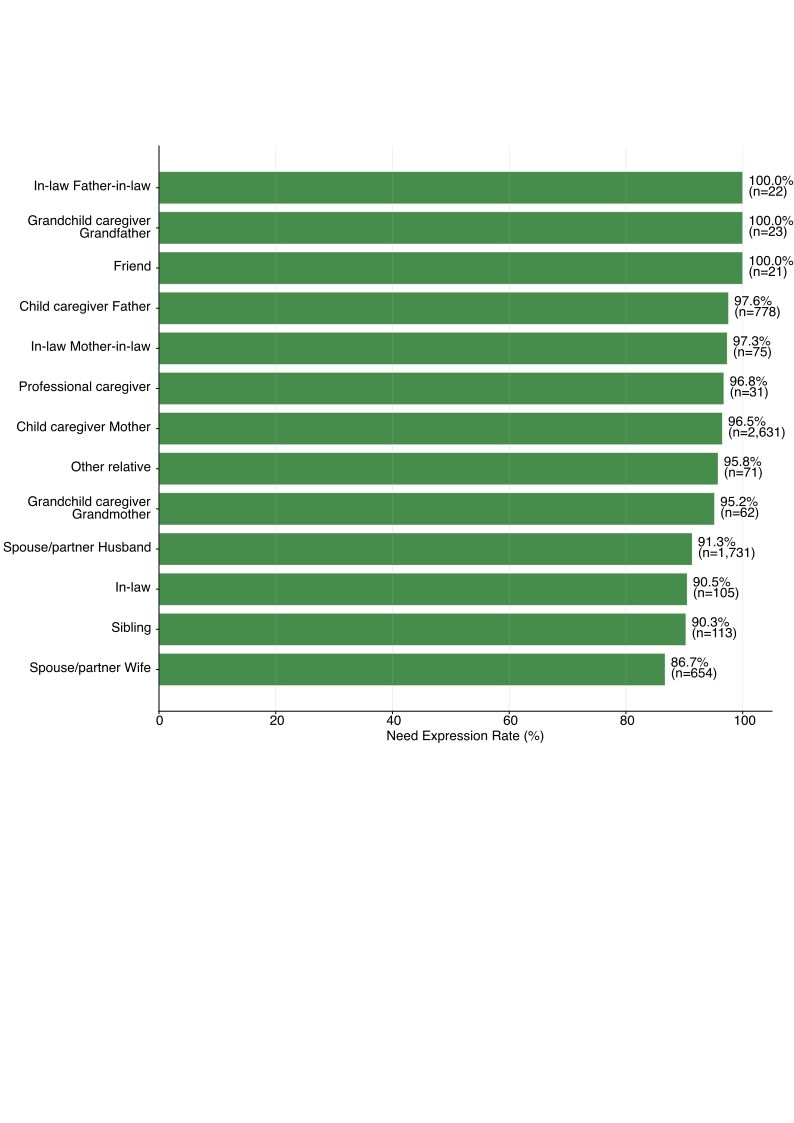

Figure S3. Overall unmet need expression by granular caregiver relationship category.

Overall unmet need expression rates are shown for each caregiver relationship subgroup. Unmet need expression was defined as the presence of any unmet need. Only relationship categories with n ≥ 20 posts are displayed. Granular categories classified as self, uncertain, or other were excluded. Percentages indicate the proportion of posts within each relationship category expressing at least one unmet need, and numbers in parentheses indicate the number of posts in each subgroup.

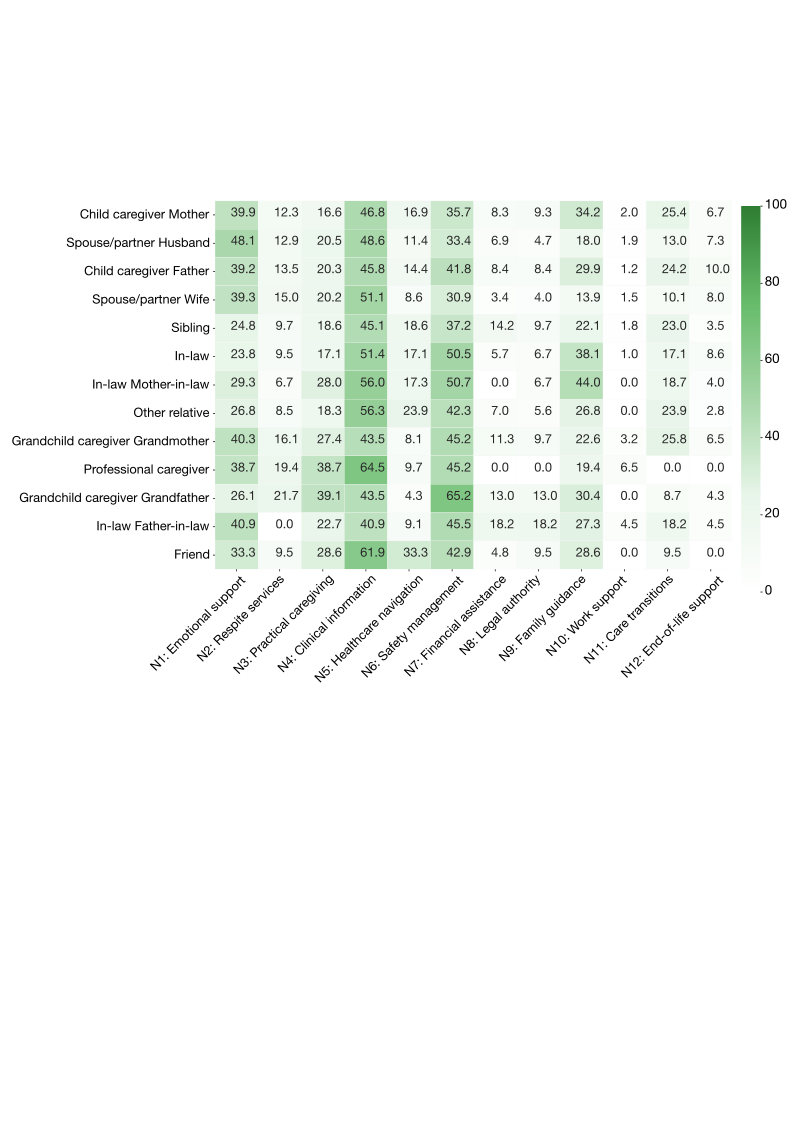

Figure S4. Unmet need category expression by granular caregiver relationship category.

Heatmap showing the percentage of posts within each caregiver relationship subgroup expressing each unmet need category. Percentages are calculated among posts within each relationship subgroup. Darker shading indicates a higher proportion of posts expressing the corresponding unmet need category. Only relationship categories with n ≥ 20 posts are shown. Granular categories classified as self, uncertain, or other were excluded. Unmet need categories include emotional support (N1), respite services (N2), practical caregiving (N3), clinical information (N4), healthcare navigation (N5), safety management (N6), financial assistance (N7), legal authority (N8), family guidance (N9), work support (N10), care transitions (N11), and end-of-life support (N12).

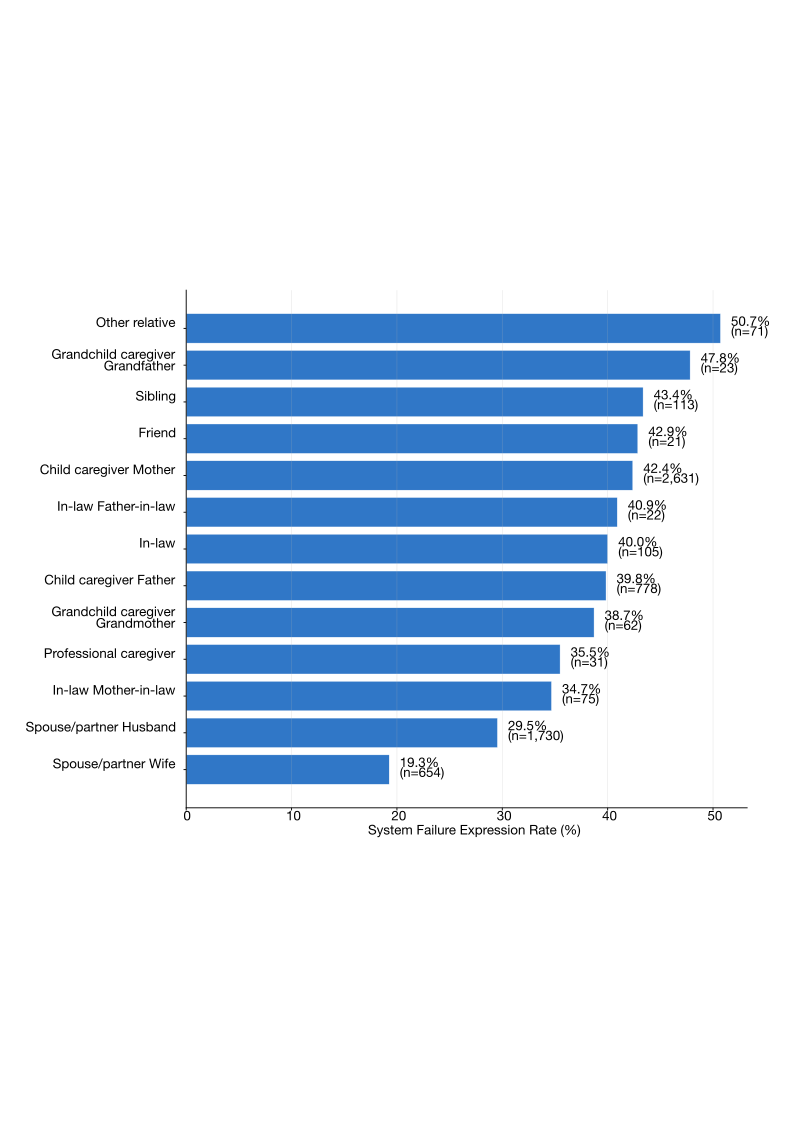

Figure S5. Overall system failure expression by granular caregiver relationship category.

Overall system failure expression rates are shown for each caregiver relationship subgroup. System failure expression was defined as the presence of any system failure. Only relationship categories with n ≥ 20 posts are displayed. Granular categories classified as self, uncertain, or other were excluded. Percentages indicate the proportion of posts within each relationship category expressing at least one system failure, and numbers in parentheses indicate the number of posts in each subgroup.

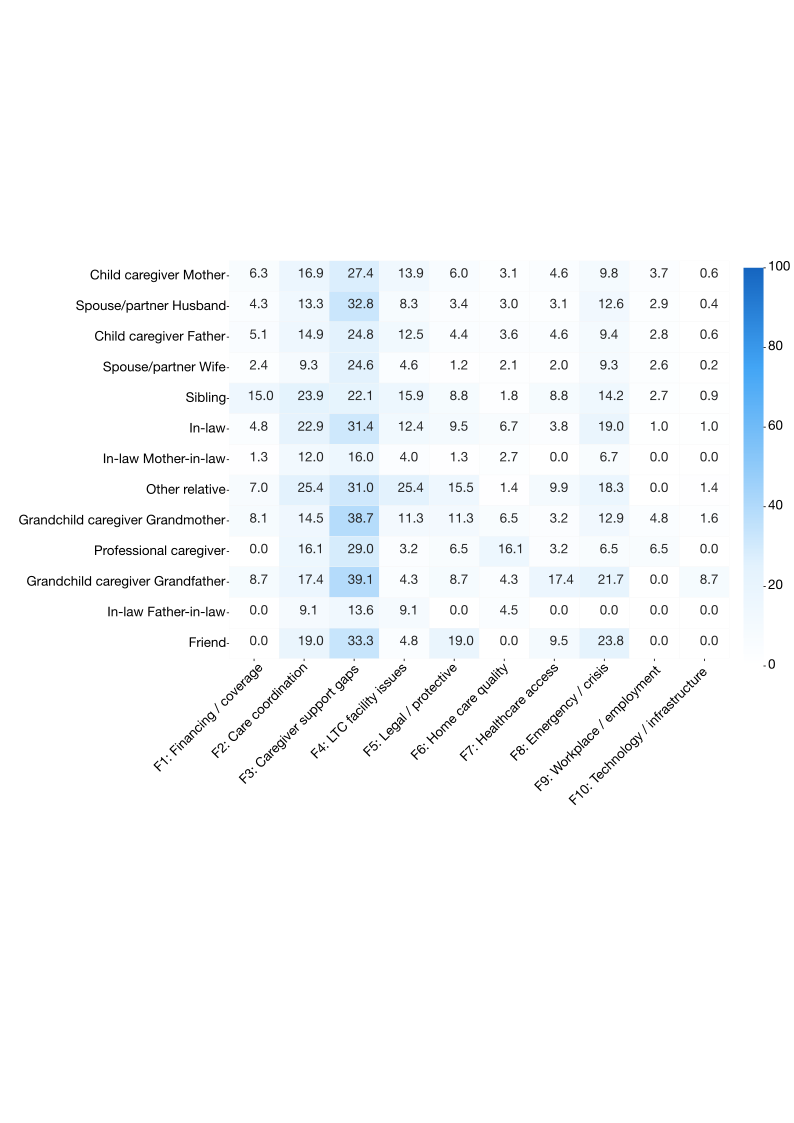

Figure S6. System failure category expression by granular caregiver relationship category.

Heatmap showing the percentage of posts within each caregiver relationship subgroup expressing each system failure category. Percentages are calculated among posts within each relationship subgroup. Darker shading indicates a higher proportion of posts expressing the corresponding system failure category. Only relationship categories with n ≥ 20 posts are shown. Granular categories classified as self, uncertain, or other were excluded. System failure categories include financing and coverage (F1), care coordination (F2), caregiver support gaps (F3), long-term care facility issues (F4), legal or protective failures (F5), home care quality (F6), healthcare access (F7), emergency or crisis-related failures (F8), workplace or employment support failures (F9), and technology or infrastructure failures (F10).
